# A nighttime telemedicine and medication delivery service to avert pediatric emergencies: An exploratory cost-effectiveness analysis

**DOI:** 10.1101/2021.09.26.21264144

**Authors:** Katelyn E. Flaherty, Molly B. Klarman, Youseline Cajusma, Justin Schon, Lerby Exantus, Valery M Beau de Rochars, Chantale Baril, Torben K. Becker, Eric J Nelson

## Abstract

**Objective:** We sought to compare the costs of a nighttime pre-emergency pediatric telemedicine and medication delivery service (TMDS) per disability-adjusted life year (DALY) averted to the costs of hospital emergency medicine (HEM) per DALY averted from a societal perspective.

**Methods:** We studied a nighttime pediatric TMDS and HEM in a semi-urban and rural region of Haiti. Costs of the 2 services were enumerated to represent the financial investments of both providers and patients. DALYs averted were calculated to represent the ‘years lives lost’ (YLL) and ‘years lost to disability’ (YLD) from diarrheal, respiratory, and skin (bacterial and scabies etiologies) disease among children from zero to 9 years old. The incremental cost-effectiveness ratio (ICER) was estimated and compared to the per-capita gross domestic product (GDP) of Haiti ($1,177). Cost-effectiveness was defined as an ICER less than 3 times the per-capita GDP of Haiti ($3,531). Univariate sensitivity analysis was performed to evaluate how uncertainty of individual parameter estimates (utilization rates, costs, lost wages, discounting factor) affected the ICER.

**Results:** The total costs of the nighttime TMDS and HEM to society were $285,931.72 per year and $89,335.41 per year, respectively. The DALYs averted by the TMDS and HEM were 199.76 and 22.37, respectively. Through sensitivity analyses, the ICER of the TMDS ranged from $791.43 to $1,593.35.

**Conclusion:** A nighttime pediatric TMDS is a cost-effective alternative to HEM for pre-emergency pediatric care in semi-urban and rural regions in Haiti, and possibly in similar lower-middle income countries.

## INTRODUCTION

Respiratory infections and diarrheal disease are the leading causes of pediatric mortality for children between one month to 5 years of age globally and resulted in 560,000 deaths in 2018.^1, 2^ Under 5 mortality (U5M) disproportionately burdens those living in low- and middle-income countries (LMIC).^3^ LMIC endure U5M rates 14 times higher than high income countries (HIC), and within LMIC, the U5M rate among the lowest wealth quintile is 2 times that of the highest quintile.^4^ The COVID-19 pandemic has increased this disparity.^5, 6^

Well-established low-cost treatments exist for both acute respiratory infections and diarrheal diseases.^7, 8^ Oral amoxicillin for bacterial pneumonia can reduce mortality by 32%^9^ and treatment of diarrhea with oral rehydration solution and zinc can reduce mortality by 93%^10^ and 23%,^11^ respectively. However, these treatments are most effective when administered soon after symptoms start, which is difficult when healthcare access is limited, especially at night. Delayed treatment, especially in children, can result in rapid progression to an emergent state.^12^ In these settings, emergency care is considerably more difficult to access and expensive than pre-emergency care.^13, 14^ More than half of the 5.3 million early childhood deaths in 2018 were considered preventable with basic healthcare.^15^

Improving access to healthcare is one of the highest global health priorities set by the Sustainable Development Goals (SDG).^16, 17^ SDG 3.8 seeks to “achieve universal health coverage”, however progress is insufficient to reach this target by 2030.^17, 18^ LMIC are the furthest off this target.^19^ Innovative, cost-effective approaches are needed to overcome persistent as well as emerging barriers to improve and sustain healthcare access in LMIC. In Haiti, only 23% of the total population and 5% of the rural population have access to quality primary care.^20^ Access to emergency services is low with 51% of the population living within 50 km of a tertiary care facility.^13^ Child health and wellness is substandard, with Haiti ranking 151 out of 180 countries on the Flourishing Index.^21^ The U5M rate is 65 per 1,000 live births compared to 39 per 1,000 live births globally.^1^ Respiratory infection and diarrheal disease are major contributors to this burden, accounting for over half of hospitalizations and over a third of deaths of hospitalized children.^22^

To better understand barriers to pediatric healthcare in Haiti, we conducted a needs assessment to identify differences in household healthcare seeking intention (‘would do’) and behavior (‘did do’);^23^ this initiative was part of the Improving Nighttime Access to Care and Treatment Study (INACT-1). In this needs assessment, households expressed an intention to seek care from a provider within conventional networks, but because of cost, distance, and nighttime hours, in practice sought care from disconnected providers near to the household. INACT-1 revealed that a pediatric telemedicine and medication delivery service (TMDS), known as MotoMeds, might provide an innovative solution to one aspect of the crisis of limited access to pediatric healthcare.

The TMDS was launched in Gressier, Haiti as a pre-pilot (INACT-2) in September 2019. Families with children experiencing pre-emergent medical problems at night called the call center. A nurse with physician oversight used a clinical decision-support tool adapted from the World Health Organization (WHO) Integrated Management of Childhood Illness guidelines^24, 25^ to triage, conduct an assessment and generate a plan and logistics system to enable household medication delivery. Patients identified with life-threatening conditions were referred for emergency care. During INACT-2, call-center nurses were dispatched to all households to confirm that call-center findings matched in-person findings (Figure 1). Respiratory illness, diarrhea and skin problems (bacterial infections, scabies) were common, and the median time to delivery was 1h 20 min. At 10-days, 94% of parents reported the problem resolved/had improved.

**Figure 1.**
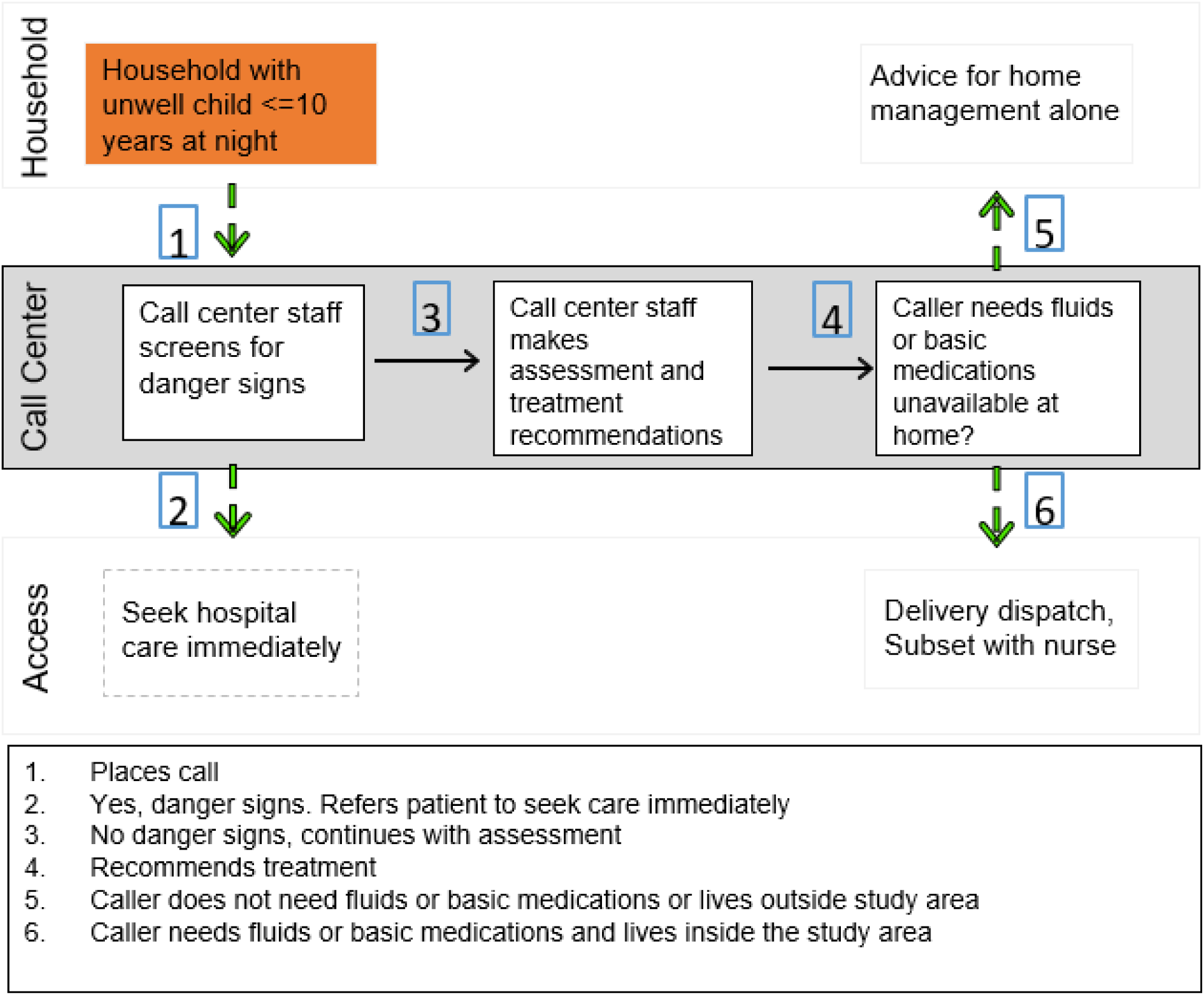
Scaled MotoMeds workflow.

The objective of the study herein was to conduct a cost-effectiveness analysis of a scaled TDMS model. We determined and compared the cost of the nighttime pre-emergency pediatric TMDS per DALY averted to HEM per DALY averted.

## METHODS

### Ethics Statement

Ethical approvals were obtained from the Comité National de Bioéthique (National Bioethics Committee of Haiti) and the University of Florida Institutional Review Board for participants in INACT-1 (1718-35; IRB201703246) and INACT-2 (1920-42; IRB201802920). For both studies, participants provided written informed consent/assent. For INACT-2, a waiver of documentation of written consent was obtained for those without a household delivery.

### Setting and participants

In Haiti, the median age is 22 years, and the life expectancy is 64.^26^ The leading causes of death across all ages are stroke, lower respiratory infections, and HIV/AIDS. Among children less than 5 years of age, the leading causes of death are acute respiratory infections, prematurity, and diarrhea.^13^ About 62% of the population lives below the international poverty level of $1.25/day.^14^ Government healthcare spending per capita is $8, below the low income average of $9.^27^ Over one third of this funding is spent at the hospital level, leading to an over reliance on emergency services.^28^ In 2019, the MotoMeds TMDS was launched in Gressier, Haiti (INACT-2), a mountainous commune of the Port-au-Prince Arrondissement with a population of 38,092.^29^ A 78 sq km delivery zone extending 5 km out from the call center was established, encompassing approximately 89% of the population. MotoMeds TMDS will expand to the similar but geographically distinct communes of Leogane and Jacmel (Figure 2) which have populations of 208,799 and 195,674 respectively.^29^ Analyses herein used data from the INACT-1 needs assessment and INACT-2 pre-pilot to evaluate the cost effectiveness of the scaled TMDS in Gressier, Leogane, and Jacmel (Figure 2). The ‘scaled’ model for MotoMeds restricts household visits to mainly moderate cases and few mild cases; severe cases bypass MotoMeds for hospital referral.

**Figure 2.**
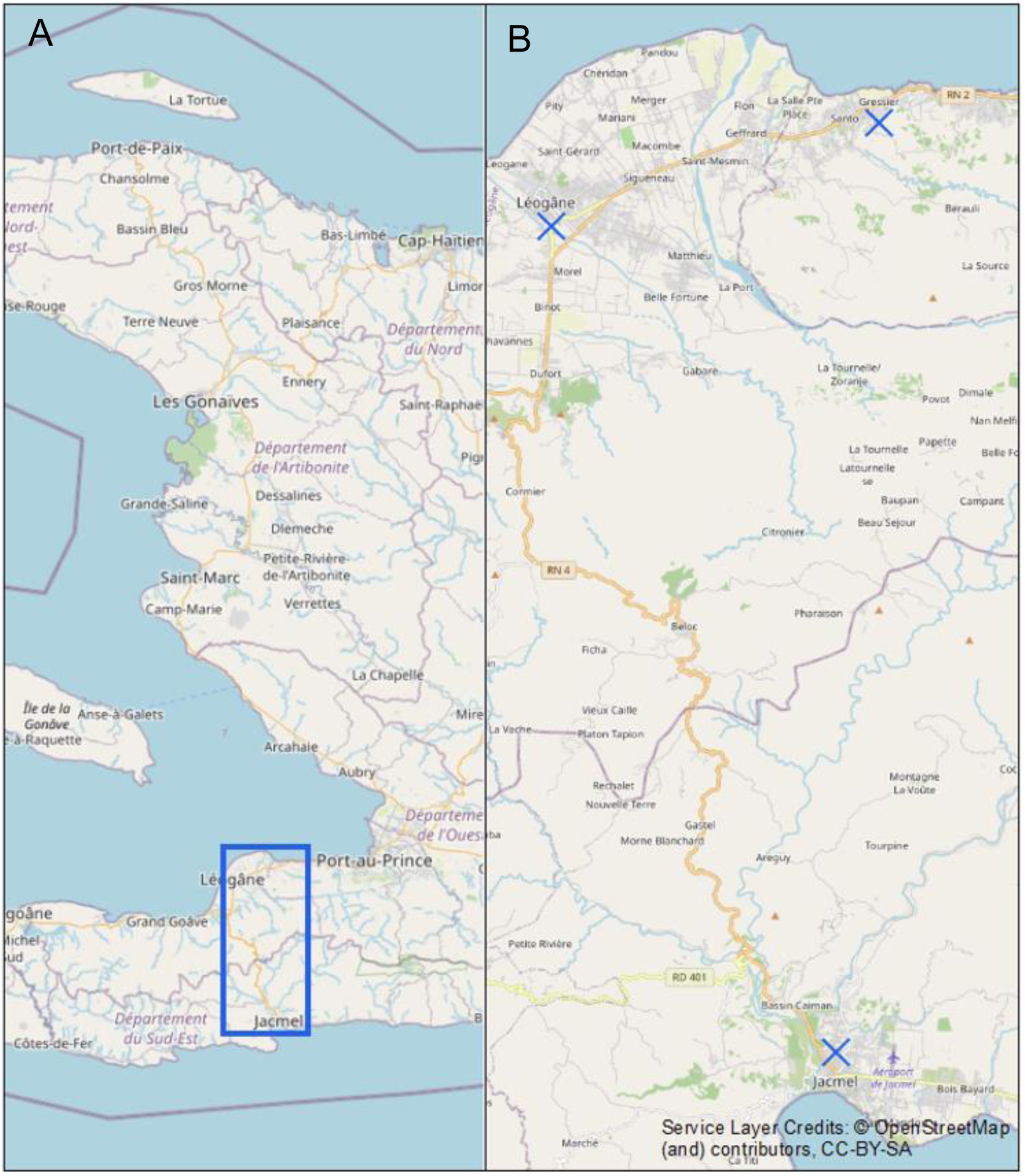
A. Map indicating extend of panel B. B. Map of MotoMeds service areas in Haiti: Gressier, Leogane, and Jacmel. Blue crosses represent MotoMeds service areas. Geospatial analyses were conducted in ArcGIS 10.7.1; basemap attributions are watermarked on figures by ArcGIS per ESRI policy^48, 49^.

### Study Design

The cost of the scaled nighttime pre-emergency pediatric TMDS per DALY averted was compared to the cost of HEM per DALY averted from a societal perspective. The rationale was that the TMDS and HEM are the only accessible healthcare options at night. Costs represent the financial investments of both providers and patients. The incremental cost-effectiveness ratio (ICER) describing additional costs of the TMDS per DALY averted was estimated and compared to the per-capita GDP of Haiti ($1,177 in 2020).^30, 31^

### Population and Utilization Estimations

Delivery zones were delineated in each location based on distance and terrain. Inhabitants within these zones were referred to as the ‘accessible population’. The delivery zone for Gressier remained unchanged from INACT-2 and encompasses the second and third communal sections. Accessible populations of Leogane encompass the first, second and third communal sections while accessible populations in Jacmel encompass the urban and half of the rural communal sections of Bas Cap Rouge, Ravine Normande and Gaillard. Populations were extrapolated from 2015 data with an estimated 4.50% growth rate to 2020.^32^ The TMDS coverage and patient load were calculated through extrapolation of INACT-2 data to account for increased advertising and awareness of the service and expansion to accessible regions of Leogane and Jacmel. HEM utilization and patient load were estimated from INACT-1 data and 2016 Demographic and Health Surveys (DHS) data relating to the proportion of the population that seeks hospital level care. INACT-1 found that families sought care for 32% of pediatric health events.^23^ The 2016 DHS in Haiti found that children under 5 sought care for 40.4% of acute respiratory infections and 32.9% of diarrheal cases.^33^ These figures were averaged to estimate 35.1% of families with a sick child seek care. INACT-1 found that 8% of care-seekers seek hospital care. DHS data were used to determine the proportion of the accessible population ages under 1 year (2.2%), from one to 4 years (7.5%), and 5 to 9 years (11.3%).^33^ The age distribution was assumed to be geographically uniform across Haiti.

### Cost Estimation

All costs were converted from Haitian gourdes to US dollars ($) at a rate of 1 US dollar to 90.15 Haitian gourdes (24-month average July 2019 to June 2021).^34^

#### Telemedicine and Delivery Service

Fixed, variable, and family costs were summed to determine the societal cost of the TMDS. Costs represented the operational budget and did not include start-up expenses. Primary data were obtained from 16 months of MotoMeds operations, the TMDS service in Gressier (INACT-2). Fixed costs refer to costs that did not vary according to patient volume while variable costs refer to costs that did vary according to patient volume. Family costs refer to the amount paid by the family for the service (Figure 3).

**Figure 3.**
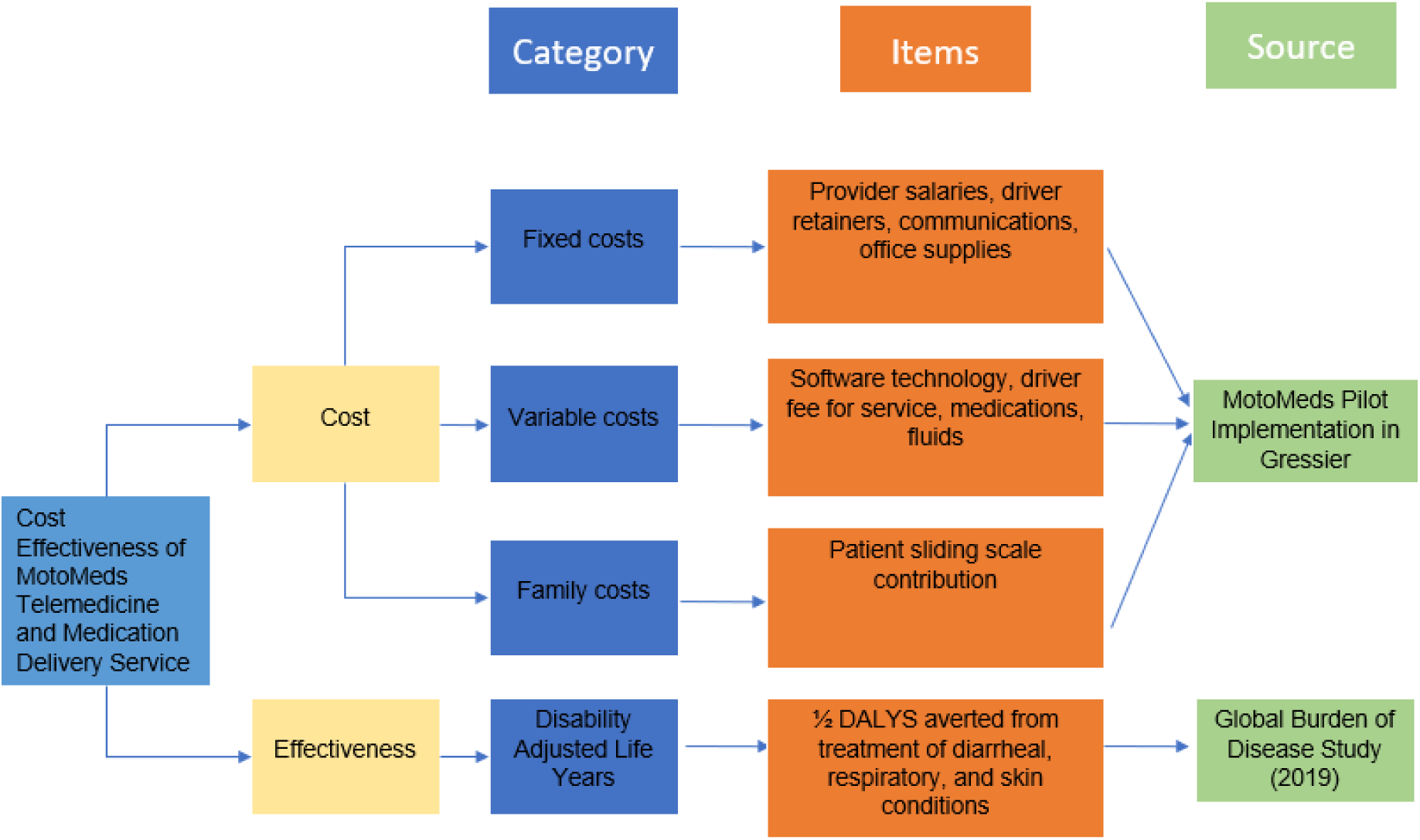
Costs and effects of MotoMeds telemedicine and medication delivery service.

##### Fixed costs

The supervisor, study physician(s), and on-call physician(s) were assumed to be paid a fixed monthly rate. The call center nurses were assumed to be compensated per shift worked. In INACT-2, Gressier drivers were paid a daily retainer; however, this was not included in the compensation scheme for Jacmel and Leogane. For the pre-pilot, the TMDS employed 2 call center nurses and 2 drivers per night. In order to serve Gressier, Leogane and Jacmel, the TMDS at scale was expected to employ 4 call center nurses, 5 on-call nurses and 14 drivers per night. Human resource administrative fees were estimated to be between 8-10%. Fixed costs also captured monthly operational costs (e.g. office rent, internet, and phone plans); drivers and on-call nurses were determined to receive a set amount of cellular phone credits monthly.

##### Variable costs

Variable costs included driver delivery payment, medications, and fluids. They were calculated per patient and multiplied by the expected number of patients per year. Half of deliveries were estimated to require 2 drivers and 20% of deliveries were estimated to require a nurse visit due to driver safety and patient severity respectively. Variable costs also captured software technologies utilized by TMDS for call intake (Twilio)^35^ and driver dispatch (Beacon by TrekMedics).^36^

##### Participant costs

Costs to participants refer to the amount families pay to use the TMDS. Under the TMDS model, families are asked to pay what they are able to on a sliding scale starting from 500 gourdes (5.78) down to zero gourdes. During INACT-2, 30% of families paid in full and 25% paid in part (250 gourdes, 2.89).^23^ We conservatively assumed the same ratio for these analyses.

#### Hospital Emergency Medicine

Patient and provider/institutional costs were summed to determine the societal cost of HEM (Figure 4). Primary data were obtained from fees at 2 main local hospitals, one private and the other public/charity, and INACT-1 data. Patient costs captured the amount families pay for pediatric care for diarrheal, respiratory, or bacterial skin disease. Patient costs associated with diarrheal disease were calculated from the costs of admission, a 2-day hospital stay, urine and stool testing (part of the local standard of practice), amoxicillin, IV fluids and oral rehydration solution. Patient costs associated with respiratory disease were calculated from the costs of admission, a 2-day hospital stay, oxygen, a hemogram, amoxicillin, and an X-ray. Patient costs associated with bacterial skin disease were determined from a single case from INACT-2 in which a pediatric patient with a bacterial skin infection required a 2-day hospital stay. Patient healthcare costs were calculated as a weighted average of the costs to treat diarrheal, respiratory, and skin disease based on condition prevalence in Haiti.^37^ Patient costs additionally captured the amount families pay for transportation to the hospital. Patient transportation costs were calculated as a weighted average of reported transportation costs from INACT-1 data and an INACT-2 bacterial skin infection case. Patient indirect costs accounted for 2 days of lost wages by a caretaker. Daily wages were estimated from the mean per capita gross national income. We were not able to obtain provider/institutional costs, likely due to the absence of specific costing data at local hospitals.^38^ As a result, provider costs were conservatively assumed to be equal to or greater than the direct healthcare patient costs for HEM.

**Figure 4.**
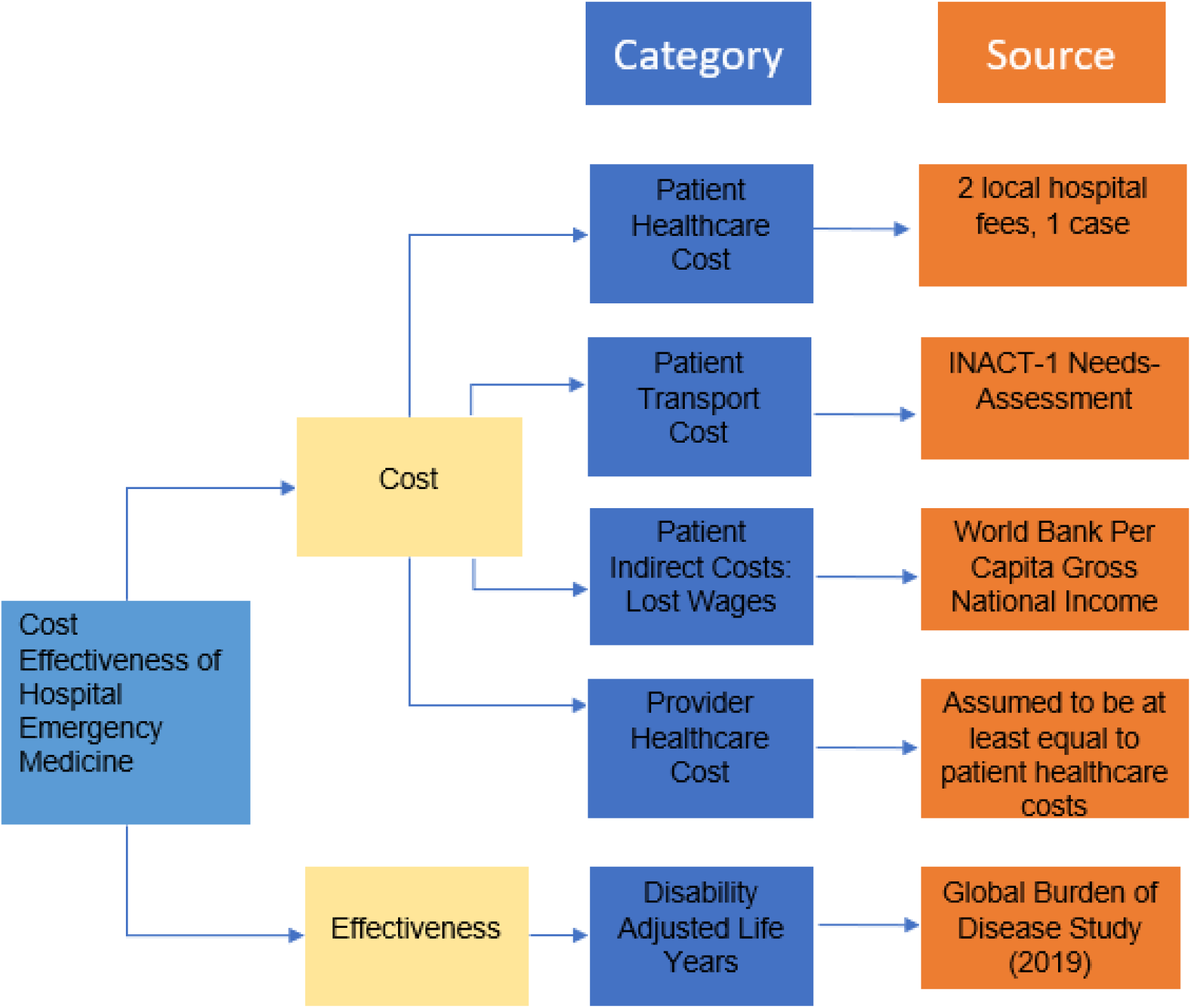
Costs and effects of hospital emergency medicine.

### Effect Estimation

#### Telemedicine Delivery Service

The key outcome indicator was the disability-adjusted life years (DALYs) averted by treatment of pediatric pre-emergency conditions. The total number of DALYs averted by a scaled TMDS were calculated using data from the 2019 Global Burden of Disease (GBD) Study^39^ to determine the years of life lost due to premature mortality (YLL) and years of healthy life lost due to disability (YLD) for the 4 conditions most commonly treated by the TMDS: respiratory infection, diarrheal disease, bacterial skin infection, and scabies skin infestation.

The total number of DALYs averted by the TMDS within the accessible population who are likely to use the TMDS for children under ten years old was estimated using the following formula: DALY = YLL + YLD. YLL was estimated by multiplying the sum of half of the mortality rates associated with respiratory infection, diarrheal disease, bacterial skin infection, and scabies skin infestation by the life expectancy for children under one year old, one to 4 years old and 5 to 9 years old, which was 63.71 years, 61.21 years and 56.48 years, respectively^40^. Mortality rates were halved to account for the TMDS operating at nighttime alone. YLD was estimated by summing half the prevalence of respiratory infection, diarrheal disease, bacterial skin infection, and scabies skin infestation multiplied by the corresponding disability weights for children aged under one year old, one to 4 years old and 5 to 9 years old per the GBD Study.^39^ Prevalence rates were halved to account for TMDS operating at nighttime only. DALY estimates were calculated for the proportion of the accessible population likely to utilize TMDS (see utilization estimates). DALY estimates were discounted at 3% in accordance with global health convention.^41^

#### Hospital Emergency Medicine

The total number of DALYs averted by patient utilization of HEM was calculated for the same 4 conditions. In YLL and YLD calculations, the full mortality and prevalence estimates were used as HEM is available at all times of day and night. DALY estimates were calculated for the proportion of the accessible population likely to utilize HEM (see utilization estimates). DALY estimates were again discounted at 3%.

### Cost-Effectiveness Estimation

The ICER was estimated using the following formula: (Cost of TMDS-Cost of HEM) / (DALYs Averted by TMDS-DALYs Averted by HEM). In accordance with WHO recommendations^42^, cost-effectiveness is defined herein as when the incremental cost-effectiveness ratio of an intervention is less than 3 times ($3,531) the per-capita GDP of Haiti ($1,177 in 2019).^31^

#### Sensitivity Analysis

Univariate sensitivity analysis was performed to evaluate how uncertainty of the parameter estimates affects the ICER. The following parameters were varied to low and high values: proportion of the population utilizing TMDS (25-75%), proportion of the population utilizing HEM (0-5%), cost of HEM care per patient ($100-180), cost of hospital transport per patient ($0-$9), cost of one day of lost wages ($0-7) and discounting rate (0-6%). The number of patients served by TMDS was varied in accordance with the proportion of the population utilizing TMDS. Likewise, number of patients served by HEM was varied in correspondence with the proportion of the population utilizing HEM.

#### Data Analysis

Data analysis was conducted in Microsoft Excel and R Version 4.0.3.^43^ DALY calculations were performed within the WHO Human Services Handbook Excel template for DALY calculation. To estimate the cost and effect of TMDS at-scale we first assumed that diarrheal disease, respiratory infections, bacterial skin infections, and scabies skin infestation will remain the conditions most commonly treated by future iterations of the TMDS. Additionally, it was assumed that TMDS will serve similar population proportions in Gressier, Leogane, and Jacmel. Furthermore, it was assumed that the exchange rate will remain relatively stable at the 24-month average.^34^

## RESULTS

### Population and Utilization Estimations

The accessible population is estimated to be 33,971 in Gressier, 173,838 in Leogane and 65,630 in Jacmel. The total accessible population is estimated to be 273,440 with 57,422 children under 10 years of age. During the INACT-2 study that had minimal advertising, one to 2 patients per night were observed. We estimate approximately 25% of the accessible population of Gressier was aware of TMDS. In a scaled TDMS model, advertising efforts are expected to be doubled to increase the utilization to 50%. This is expected to result in 4 patients per night in Gressier,18 patients per night in Leogane, and 7 patients per night in Jacmel, in correspondence with the accessible populations of each location. Therefore, a scaled TMDS model is anticipated to serve 29 patients per night and 10,585 patients per year. Given 35% of families with a sick child seek care and 8% of care-seekers seek hospital care, it is estimated that 2.8% of the accessible population will utilize HEM, 47.2% less than TMDS, for a total of 296 cases per year (Supplement 1).

### Cost Estimations

#### Telemedicine and Medication Delivery Service

Fixed costs totaled $105,530.97 while variable costs totaled $155,446.61 for 10,585 patient encounters per year. Patient costs were estimated at $24,954.14. Thus, assuming 10,585 patients encounters per year by the TDMS, the total cost of the TMDS to society is estimated at $285,931.72 per year (Table 1). The costliest component of the TMDS is staff compensation, specifically nurse and driver compensation.

**Table 1.** MotoMeds Yearly Societal Costs

| **Fixed Costs** | Amount | Notes |
| --- | --- | --- |
| Clinical Staff | \$82,627.24 | Physician and nurse salaries |
| Non-Clinical Staff | \$5,975.05 | Director salary, driver retainers |
| Office | \$8,088.41 | Rent, supplies |
| Communication | \$6,440.28 | Phone, internet |
| Advertising | \$2,400.00 | Flyers, radio time |
| **Variable Costs** | Amount | Notes |
| Driver | \$112,762.01 | Compensation for deliveries with and without nurse |
| Treatment | \$33,448.60 | Medications, fluids |
| Technology | \$9,236.00 | Twilio, Beacon |
| **Patient Costs** | Amount | Notes |
| Family Payment | \$24,954.14 | Family full or partial payment |
| **Total Societal Cost:** | **\$285,931.72** | |

#### Hospital Emergency Medicine

The estimated patient cost of HEM for pneumonia was $200.78 at one local emergency center and $151.41 at another local emergency center for an average of $176.10. The estimated patient cost of HEM for diarrhea was $130.62 one center and $59.62 at the other for an average of $95.12. The patient cost of HEM for a bacterial skin infection was $150.03 at one local emergency center. Therefore, when averaged using global burden of disease prevalence weights, the estimated patient cost of HEM is $144.77 per patient.^39^ In INACT-1, 9 pediatric patients who sought HEM reported an average transportation cost of $4.13 (daytime vs nighttime was not known). In INACT-2, a single patient reported a transportation cost of $8.87 at night. Using these figures the weighted average estimate for hospital transportation cost is $4.60 per patient. The 2019 per capita gross national income in Haiti was reported at $1,330/year ($3.64/day)^44^. As a result, indirect costs include $7.28 in care giver lost wages per patient for 2 days of HEM. Provider/institutional costs were conservatively assumed to be equal to or greater than the direct patient healthcare costs of $144.77 per patient. Assuming 296 patients access HEM per year, 2.8% of the accessible population, the total cost of HEM to society is $89,335.41 (Table 1).

### Effect Estimations

#### Telemedicine and Medication Delivery Service

Nighttime TMDS treatment for diarrheal diseases, respiratory infections, bacterial skin infections, and scabies skin infestations was found to avert 17.1 YLL and 28.6 YLD among children under 1 year old, 25.1 YLL and 68.2 YLD among children one to 4 years old, and 4.3 YLL and 56.4 YLD among children 5 to 9 years old. It was determined that the TMDS would avert 199.76 DALYs per year of operation.

#### Hospital Emergency Medicine

HEM treatment of diarrheal diseases, respiratory infections, bacterial skin infections, and scabies skin infestations was found to avert 1.9 years of life lost and 3.2 years lost to disability among children aged 0, 2.8 YLL and 7.6 YLD among children 1 to 4, and 0.5 YLL and 6.3 YLD among children 5 to 9. It was determined that HEM would avert 22.37 DALYs per year.

In the TMDS and HEM DALY calculations, treatment of respiratory infections was found to avert the most YLL in all age groups. Treatment of diarrheal disease was found to avert the most YLD in all age groups.

##### Cost-Effectiveness Estimation

The TMDS is estimated to cost society $196,596.31 more than HEM yet averts an additional 177.39 DALYs. Correspondingly, the ICER is estimated at $1,108.27 signifying the TMDS costs an additional $1,108.27 to avert one additional DALY. This value is under the per capita GDP of Haiti, $1,177 in 2019,^31^ therefore the TMDS is considered highly cost effective by WHO standards.^30^

##### Sensitivity Analysis

Through sensitivity analysis the ICER ranged from $791.43 to $1,593 (Figure 5). When the following parameters were increased, TMDS utilization, HEM utilization, cost of HEM care, cost of hospital transport and daily wages, the ICER decreased, signifying a higher degree of cost-effectiveness. A decreased discounting factor was likewise associated with a lower ICER signifying a higher degree of cost-effectiveness. Through all parameter variations the ICER remained within 2x the per capita GDP of Haiti and thus signified cost-effectiveness.

**Figure 5.**
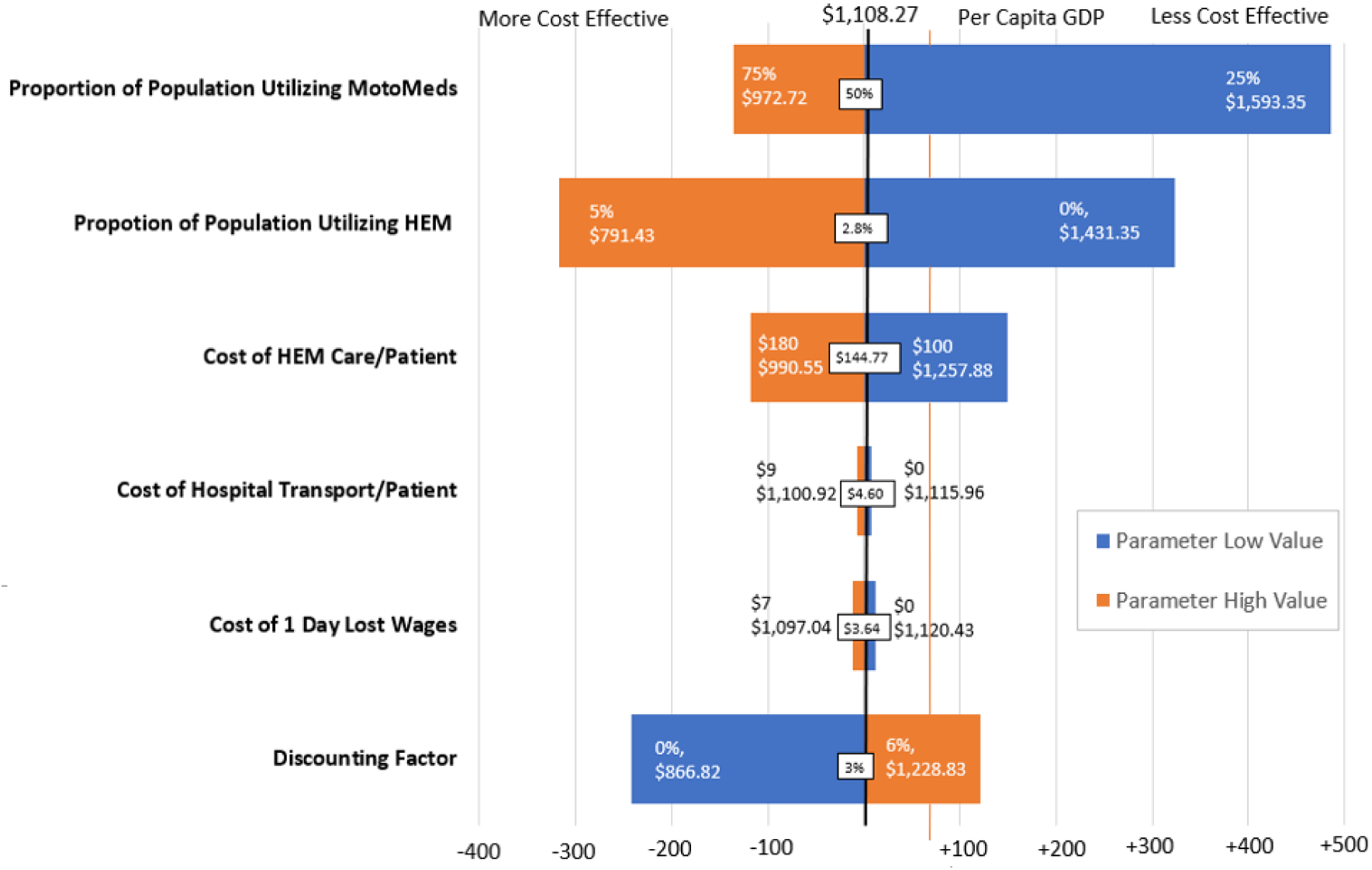
Sensitivity Analysis. Univariate sensitivity analysis was performed to evaluate how uncertainty of individual parameter estimates affected the Incremental Cost Effectiveness Ratio in relation to the per capita GDP in Haiti. HEM = hospital emergency medicine.

## DISCUSSION

Here, we demonstrated the cost-effectiveness of a nighttime TMDS compared to hospital emergency services in Haiti. TMDS has the potential to avert additional years of life lost and years lost to disability among children under 10 for a relatively low cost to society. TMDS greatest costs relate to compensation of drivers and nurses. In this way, finances ideally are funneled directly back into society.

The results were robust to all input variables. Sensitivity analyses showed that the proportion of the population utilizing TMDS and the proportion of the population utilizing HEM were the key parameters influencing the ICER (Figure 5). Increased TMDS utilization and increased HEM utilization are positively associated with the ICER.

While alternative TMDS operational modes and scales were not considered, it is important to note that increased TMDS utilization was found to be positively associated with cost-effectiveness. Therefore, a positive feedback loop where professionalism, reasonable payment structure and word-of-mouth might perpetuate TMDS usage and has the potential to further increase the cost-effectiveness of the service.

This cost-effectiveness analysis may stimulate interest in innovative solutions to improve access to care like the TMDS evaluated herein. Other TMDSs have been largely limited to HIC^45^; however there is likely an underappreciated need for both telemedicine and medication delivery in LMIC. Successful telemedicine use cases include the Aponjon remote consultation service for maternal, neonatal, and infants in Bangladesh that provided valuable medical advice and support but lacked a referral system^46^ and the Uliza! Clinicians’ HIV Hotline in Kenya that effectively assisted healthcare providers to manage their HIV+ patients^47^. Successful medication use cases include Riders for Health that champions efficient diagnosis of infectious diseases to prevent progression and spread in 7 LMIC^21^. The cost-effectiveness analysis herein may bolster support for these initiatives and other TMDS in LMIC.

Weaknesses of this study include failure to obtain the provider/institutional costs of hospital emergency care. It is likely that local hospitals do not have the level of granular data needed and efforts to find it in the literature were unsuccessful.^38^

Other weaknesses include failure to account for patients with nighttime symptoms progressing at a pace sufficiently slow to seek basic clinical care in the morning. In Haiti, primary clinical care is not available at night and thus was not included as a comparator. However, to evaluate the true cost-effectiveness of TMDS, future studies should consider the proportion of non-progressive cases and the cost of morning clinical care. Other future analyses should consider the deteriorated state of patients seeking emergency care.

Limitations in data availability likely contributed to a conservatively low ICER. The estimate of HEM usage is based on the INACT-1 study where families were asked to recall illnesses from the past month but did not specifically ask about illnesses requiring pediatric hospital level care, with the true HEM usage likely to be higher. MotoMeds TMDS cares for patients 10 years and under but DALYs in this analysis were calculated for children 0-9 years. Additionally, TMDS averts DALYs caused by other illnesses not included in this analysis.

### Conclusions

A TDMS for nighttime pre-emergency pediatric care is cost-effective compared to HEM in semi-urban and rural Haiti. This form of TDMS may represent an innovative option to extend access to care to isolated families at night across Haiti and possibly in other LMIC with similar challenges.

## Data Availability

All pertinent data is included in the manuscript.

## SUPPLEMENT

**Supplementary Table 1.**
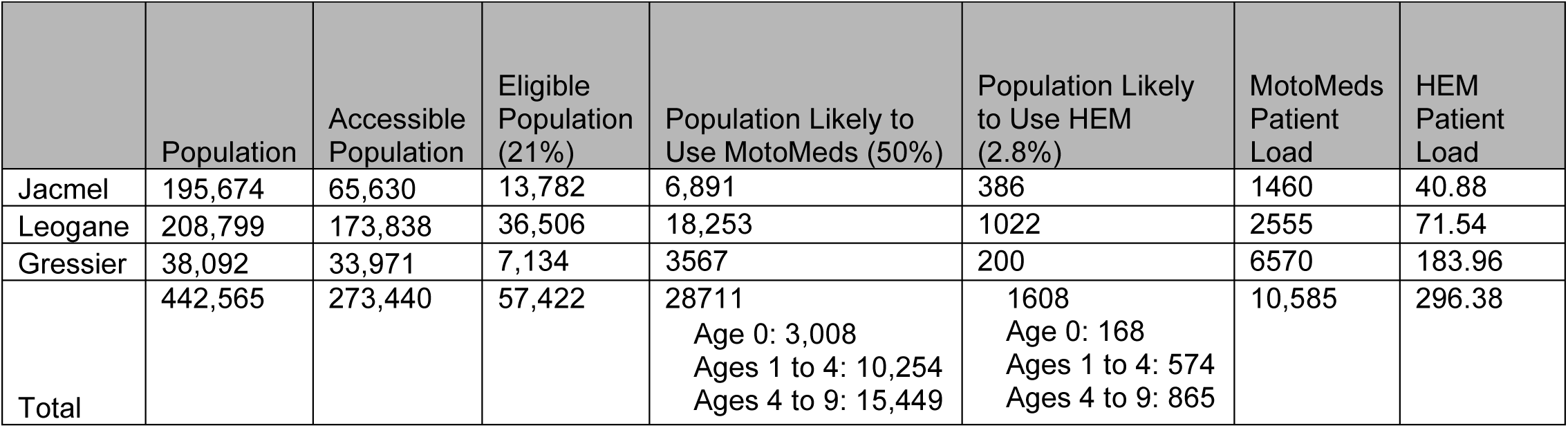
Estimates of population accessible and covered by MotoMeds

|  | Population | Accessible Population | Eligible Population (21%) | Population Likely to Use MotoMeds (50%) | Population Likely to Use HEM (2.8%) | MotoMeds Patient Load | HEM Patient Load |
| --- | --- | --- | --- | --- | --- | --- | --- |
| Jacmel | 195,674 | 65,630 | 13,782 | 6,891 | 386 | 1460 | 40.88 |
| Leogane | 208,799 | 173,838 | 36,506 | 18,253 | 1022 | 2555 | 71.54 |
| Gressier | 38,092 | 33,971 | 7,134 | 3567 | 200 | 6570 | 183.96 |
| Total | 442,565 | 273,440 | 57,422 | 28711<br>Age 0: 3,008<br>Ages 1 to 4: 10,254<br>Ages 4 to 9: 15,449 | 1608<br>Age 0: 168<br>Ages 1 to 4: 574<br>Ages 4 to 9: 865 | 10,585 | 296.38 |

## Acknowledgements

We thank the team who collected the data for this study. We are grateful to Randy Autrey and Krista Berquist for their administrative support, as well as Glenn Morris at the Emerging Pathogens Institute, Desmond Schatz in the Department of Pediatrics at the University of Florida for their ongoing support and guidance from the Ministry of Public Health and Population (Ministère de la Santé Publique et de la Population - MSPP). Map data is copyrighted by OpenStreetMap contributors and available from https://www.openstreetmap.org.

## Financial Support

This work was supported by the National Institutes of Health grants to EJN [DP5OD019893] and internal support from the Emerging Pathogens Institute (EJN), the Departments of Pediatrics (EJN), the Department of Environmental and Global Health (EJN), and the Department of Emergency Medicine (KEF/TKB) at the University of Florida.

## Disclosures

The funders had no role in study design, data collection and analysis, decision to publish, or preparation of the manuscript. All authors: No reported conflicts.

## Co-Author Contact Information

Katelyn Flaherty

Departments of Emergency Medicine and Environmental and Global Health, University of Florida, Gainesville, FL, USA

Molly Klarman

Departments of Pediatrics and Environmental and Global Health, University of Florida, Gainesville, FL, USA

Youseline Cajsuma

Departments of Pediatrics and Environmental and Global Health, University of Florida, Gainesville, FL, USA

Justin Schon

College of Arts and Sciences, College of William and Mary, Williamsburg, VA, USA

Lerby Exantus

Université d’État d’Haiti-Faculté de Médecine et de Pharmacie, Port-au-Prince, Haiti

Valery M Beau de Rochars

Department of Health Services Research, Management and Policy, College of Public Health and Health Professions, University of Florida, Gainesville, FL. US

Chantale Baril

Université d’État d’Haiti-Faculté de Médecine et de Pharmacie, Port-au-Prince, Haiti

Torben K Becker,

Departments of Emergency Medicine and Environmental and Global Health, University of Florida, Gainesville, FL, USA

Eric J Nelson

Departments of Pediatrics and Environmental and Global Health, University of Florida, Gainesville, FL, USA

## Notes

### Competing Interest Statement

The authors have declared no competing interest.

### Author Declarations

Ethical approvals were obtained from the Comite National de Bioethique (National Bioethics Committee of Haiti) and the University of Florida Institutional Review Board for participants in INACT-1 (1718-35; IRB201703246) and INACT-2 (1920-42; IRB201802920). For both studies, participants provided written informed consent/assent. For INACT-2, a waiver of documentation of written consent was obtained for those without a household delivery.

